# Factors influencing access to diabetic eye care among people living with diabetes attending a tertiary hospital in Dar es salaam, Tanzania: a qualitative study

**DOI:** 10.1101/2025.10.04.25337039

**Authors:** Henry Ngogo, Deodatus Kakoko, Thadeus Ruwaichi, Nicolaus Chaula

## Abstract

**Background:** The increase in prevalence of Diabetes has led to a rise in the burden of blindness due to diabetic retinopathy. Timely access to diabetic eye care may prevent blindness due to diabetic complications, reducing the burden of blindness in countries with limited resources like Tanzania. This study aimed to explore the factors influencing access to diabetic eye care among people living with diabetes attending a tertiary hospital.

**Methods:** A total of 14 interviews were conducted with people living with diabetes attending a tertiary hospital from March 2023 to May 2023. Participants were purposively selected during their visits to the eye clinic. The interviews were audio-recorded, transcribed verbatim, and analysed by qualitative content analysis.

**Results:** Factors influencing access to diabetic eye care among people living with diabetes were reported, including the cost of eye care services, not knowing that diabetes can affect eyes, fear, and having other priorities emerged as barriers to accessing care while referral from primary healthcare workers, eye symptoms, knowing that diabetes can affect eyes, living close to the facility offering eye care services, influence from close relatives, and having health insurance were reported as facilitators to attending diabetic eye care services.

**Conclusion:** Educating individuals with diabetes about the importance of diabetic eye care, while addressing the barriers to accessing it, will help them receive timely diabetic eye care and prevent blindness caused by diabetic retinopathy.

## Introduction

Diabetic retinopathy (DR) is the ocular complication of diabetes mellitus (DM) that affects about 1 in 3 people living with diabetes (PLWD).^1^With the projected increases in the number of people living with diabetes, especially in low and middle-income countries (LMIC), the number of people with DR will also increase,^2,3^ In Tanzania, the prevalence of DR amongst PLWD attending diabetic clinics is estimated to be about 27.9%.^4^

PLWD are recommended to undergo screening for DR, which is a cost-effective intervention for preventing irreversible visual impairment due to DR. People identified as having potentially sight-threatening DR (STDR) are then referred after screening to specialist eye clinics for assessment. Diabetic eye care services in Tanzania are mostly offered in specialist referral hospitals.^5^ However, in Tanzania, compliance with referral advice is low, with only around 40% of people referred actually attending the specialist eye clinics.^4^ Several factors have been noted to influence this behaviour, including costs of eye care services, ill health, lack of awareness, and distance to the health facility. ^5–7^

This qualitative study aimed to explore factors influencing access to diabetic eye care (from DR screening services to specialist eye care) amongst PLWD attending a tertiary hospital in Dar es salaam, Tanzania.

The study was guided by the capability, opportunity, motivation, and behaviour (COM-B) model.^8^ The COM-B model has been used in several studies to examine factors influencing health-seeking behaviour among individuals ^9–11^. This framework is comprehensive and aids in identifying factors influencing a certain behaviour under the capability, motivation, and opportunity constructs.

## Methodology

### Study design

We undertook a qualitative study using in-depth interviews to explore the factors influencing access to diabetic eye care among PLWD attending a tertiary hospital in Dar es Salaam, Tanzania.

### Study settings

The study was conducted at CCBRT, which is a zonal referral hospital that receives referrals from across Tanzania. The hospital has a retina unit that provides outpatient services, pan-retinal photocoagulation (PRP) laser, anti-vascular endothelial growth factor (Anti-VEGF) injections, and vitreoretinal surgeries. At this Tertiary Hospital, DR screening services are provided at the eye Outpatient Department (OPD) for all patients with DM attending the Hospital, and those who are diagnosed with DR are being referred to the retina unit for management.

### Study population

PLWD attending their yearly DR screening or those receiving treatment for DR. PLWD who had been diagnosed with diabetes for at least one year or more, and those who had attended eye care at least once in the past, were eligible to participate in this study.

### Sampling procedures

To obtain richly informed cases, we employed purposeful sampling to select PLWD. With the help of the nurse in charge of OPD services, we selected patients from the list of patients with appointments on a particular date and asked the patients about their willingness to participate in the study, and those who agreed to participate were interviewed after exiting the services. To understand the barriers to accessing diabetic eye care services, we recruited some participants who were late and had severe DR to understand the reasons why they came late to the hospital.

### Data collection tool

An interview guide was developed by referring to the COM-B Model and the literature^5,7,8,11–16^. The guide included questions and prompts to explore the experience of PLWD when seeking eye care.

### Data collection procedure

Data were collected by the principal investigator, aided by a research assistant who was taking notes. Data were collected through in-depth interviews and recorded using an audio recorder. Data were collected in a secure and quiet conference room to ensure privacy; data were collected from March 2023 to May 2023. All interviews were conducted in Kiswahili, a widely spoken language in Tanzania. Interviews stopped after the saturation point, where no new information was coming in, even after phrasing questions differently.

### Data processing and analysis

The interviews were transcribed verbatim, and transcripts were translated from Kiswahili to English. The English transcripts were uploaded into NVivo 12 software for data analysis. We analysed data using qualitative content analysis by starting with the start list codes from the research objectives and theoretical framework. Then we inductively analysed the transcripts to come up with more codes. By comparison, we abstracted codes into sub-categories and sub-categories into categories.

### Ethical consideration

The ethical clearance to conduct this study was granted by the Muhimbili University of Health and Allied Sciences (Ref. No.DA.282/298/01.C/1557). All participants were given consent forms to read and sign before interviews.

## RESULTS

### Participants Characteristics

We interviewed 14 patients with diabetes attending an eye clinic at CCBRT Hospital. The mean age of the participants was 52 years, ranging from 35 to 69, with equal participation of men and women. 28.6% of the participants were using cash to access care, while the remaining had health insurance. The mean time that participants lived with diabetes was 11 years, ranging from 1 to 27 years.

**Table 1.**
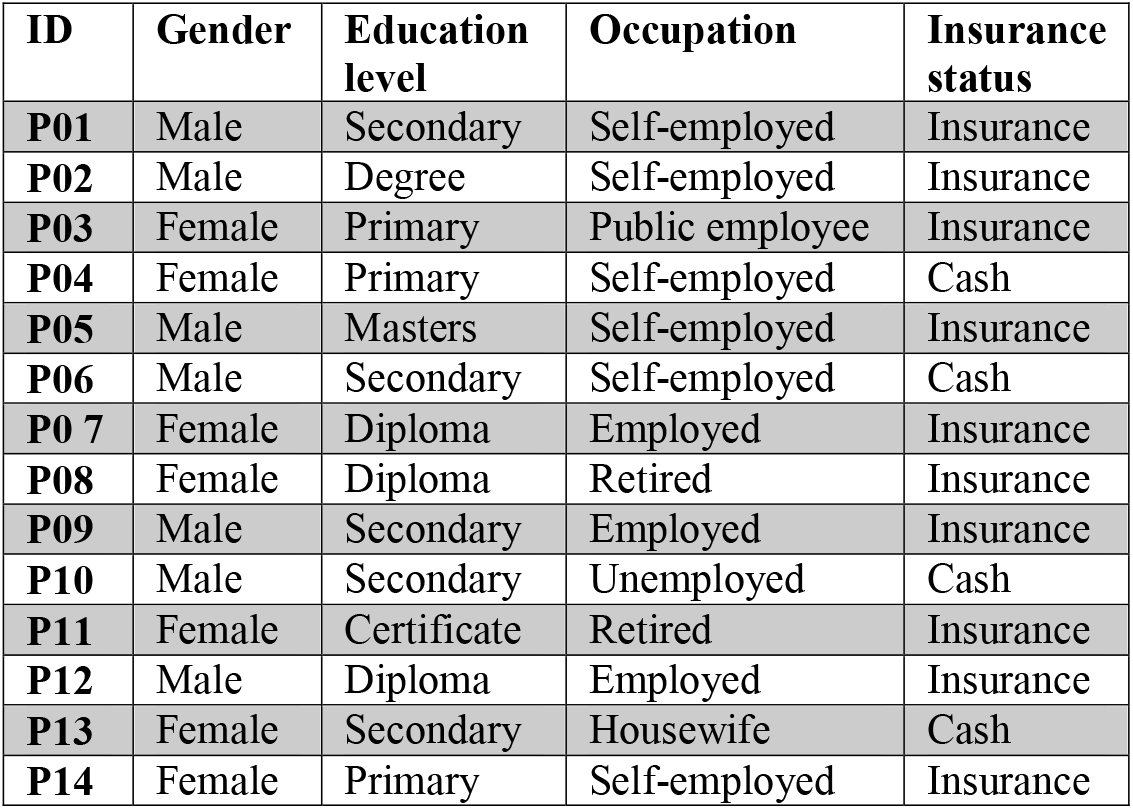
Participants’ Demographics.

| ID | Gender | Education level | Occupation | Insurance status |
| --- | --- | --- | --- | --- |
| P01 | Male | Secondary | Self-employed | Insurance |
| P02 | Male | Degree | Self-employed | Insurance |
| P03 | Female | Primary | Public employee | Insurance |
| P04 | Female | Primary | Self-employed | Cash |
| P05 | Male | Masters | Self-employed | Insurance |
| P06 | Male | Secondary | Self-employed | Cash |
| P07 | Female | Diploma | Employed | Insurance |
| P08 | Female | Diploma | Retired | Insurance |
| P09 | Male | Secondary | Employed | Insurance |
| P10 | Male | Secondary | Unemployed | Cash |
| P11 | Female | Certificate | Retired | Insurance |
| P12 | Male | Diploma | Employed | Insurance |
| P13 | Female | Secondary | Housewife | Cash |
| P14 | Female | Primary | Self-employed | Insurance |

Two broad categories that emerged during the analysis of data are Barriers to accessing diabetic eye care and Facilitators to accessing eye care.

### Barriers to accessing diabetic eye care

PLWD reported several issues limiting their attendance at the hospital for diabetic eye care services, including distance to the health facility, cost of healthcare services, and not knowing that diabetes can cause damage to their eyes.

Participants reported that they did not know that diabetes could affect their eyes; therefore, they did not appreciate the importance of attending eye care. Some of the participants reported that they sought eye care only after they began to experience problems with their vision.

> *“Initially, I did not know if there was any connection, and since I did not go for regular check-ups, I took it as normal”* (P01, Male).

Some participants reported that they did not seek timely eye care because of the fear of the unknown or because of what they would be told about their eyes and general health. However, some participants had concerns about the health care systems and quality of care, such as long hospital queues and fear that treatment options such as eye injections and laser could cause further damage to their eyes, which acted as a barrier to accessing timely diabetic eye care.

> *“You know, many people do not like getting tested; it is also fear. Fear in the sense that they believe that if they go to the hospital, there will be many people and queues”* (P01, Male)

Some PLWD reported that work, business, and home chores were barriers to accessing eye care. Participants who were business owners reported that it is difficult to leave their businesses and attend diabetic eye care when they have no one to leave their business with. While those who were employed reported that it is sometimes difficult to secure permission from work.

> *“For me personally, what hinders me the most is my business, you think about leaving your business and going away, and then you think about who will manage it in your absence”* (P01, Male)

The cost of eye care services was among the reported reasons why some participants did not access diabetic eye care services on time. Some participants stated that they had no money, and that was why they did not attend eye care services.

> *“I realised the importance of attending and continuing to attend these appointments, but I have delayed coming because I did not have the money yet”* (P14, Female).

However, some participants, especially those who were not from Dar es salaam reported indirect costs such as transport and lodging costs as the primary reason for non-attendance at their appointments.

> *“Maybe it is money, financial difficulties in the future, because even travelling from there to come here is costly”* (P10, Male).

Some PLWD reported that they were receiving routine diabetic care, and they were not told by their doctors to seek diabetic eye care; therefore, they didn’t find a reason to go, as they believed that their doctors would have told them if it was necessary. Other participants went to test their eyes for glasses but were not screened for DR, which was a missed opportunity for them to access diabetic eye care services.

> *“As I mentioned, even the doctor who spoke to me about diabetes did not tell me to have an eye examination, so there may be some disconnect between the two”* (P05, Male).

Some participants reported not going for DR follow-up visits due to sociocultural issues such as religious festivals, travelling to their homelands, taking care of the sick, or bereavement.

> *“Problems happened during Ramadan, I had difficulty coming because of Ramadan, but after Ramadan, I was able to come”* (P13, Female).

### Facilitators for accessing diabetic eye care

These factors influenced people’s access to timely diabetic eye care. These factors included knowing that diabetes can cause damage to the eyes and living near the facility offering DR services.

Participants who knew that diabetes could cause damage to their eyes reported visiting the hospital because they understood that diabetes can cause eye problems; they therefore went to the hospital to avoid such consequences. Some other participants came after seeing people close to them become visually impaired due to DM.

> *“I have heard that if blood sugar levels are too high, it can affect your vision, and I’ve heard that is why I came to test my eyes”* (P05, Male).

The need to maintain independence and career goals was the motivation behind visiting diabetic eye care follow-up for some participants, who explained that they needed to see so that they can be independent and continue to work.

> *“It is of great importance because my activities require good eyesight. Since they depend on my eyes, I prioritise taking care of them”* (P02, Male).

Some participants reported going for eye care after they experienced problems with their eyes, such as a diminution of eyesight or pain in their eyes; these symptoms made them seek eye care.

> *“Well, I started feeling some pain in my eyes, and they gave me glasses. When I wore those glasses, I felt like certain areas were completely dark that would last for a while, and then the pain slowly disappeared. Also, when I wore the glasses, I felt dizzy. When I walked, I felt dizzy and almost fell. That made me come here”* (P14, Female).

Physicians and other primary healthcare practitioners acted as important bridges between diabetic care and diabetic eye care, as some of the participants reported that they were referred for eye check-ups by their doctors managing their diabetes, while other participants reported that they were referred from optical clinics.

> *“My doctor has always advised me to check my eyes regularly; they have emphasised that diabetes is one of the things that can harm the eyes quickly”* (P02, Male)

Having a positive experience when receiving eye care was a motivating factor for participants to attend future follow-up visits, and some participants reported that they had a good experience while receiving eye care services and less waiting time, which made it easy for them to come for future visits since they were assured of good services.

> *“Yeah, I had a good experience, and I do not see any reason why I would not come back in the future. I have not encountered any issues that would make me think I do not have to come back. It has been straightforward, and even at the hospital, I found the process to be smooth, so I did not waste my time. So, I think it is good”* (P06, Male)

For those participants who were living or working nearby, the hospital offering diabetic eye care services reported that it was easy for them to attend diabetic eye care because of their proximity. Therefore, distance is among the factors that influence access to care, and those who live nearby or in Dar es Salaam reported easy access to diabetic eye care.

> *“Well, I do not think I will face any difficulties. Luckily, my office is very close to this hospital, so I will be able to come here easily and frequently”* (P05, Male).

Other participants reported they were influenced by their close relatives, who were reminding them of their diabetic eye care schedule and that they were driving them to the hospital for eye checkups, which acted as a facilitator for them to adhere to their diabetic eye care schedule.

> *“Now that I have already come here, I will keep coming here regularly because my children are here in Dar es Salaam. I will tell them to bring me here, and I will be informed that it is time for my eye check-up, so that I do not go blind. I fear going blind”* (P04, Female).

Having health insurance was a motivating factor for several participants seeking and adhering to diabetic eye care. Several participants reported that having health insurance was the reason for them to adhere to diabetic eye care, as most of the bills were covered by their insurance, making it easy for them to access timely care.

> *“No, I have not faced any difficulties because I have been using the NHIF card. Therefore, any difficulties I might face would occur if I needed to undergo tests but did not have the money, but with the NHIF card, it helps me access all the necessary tests whenever I need them”* (P02, Male)

## Discussion

In the present study, we explored the factors influencing access to diabetic eye care among PLWD. Several issues, such as not knowing that diabetes can affect eyes, the cost of eye care services, and competing priorities such as work and business, emerged as barriers to accessing eye care while living near the health facility, influence from close relatives, having health insurance and referral from other healthcare workers acted as facilitators of accessing eye care.

In this study, some participants reported that they did not know that they needed to get their eyes screened periodically, and that was the reason for not attending eye care services, as in some studies ^6,7,12^ which reported that some PLWD did not attend DR screening because they did not know they needed it. This finding shows the importance of educating PLWD about the importance of DR screening and eye care services to prevent avoidable blindness caused by DR, as knowledge about DR can positively influence PLWD to attend DR screening.^6,12^

The cost of accessing services has been reported to be a barrier to care by several studies ^5,6^, This has been reported by participants in this study, where PLWD could not access eye care services due to associated costs. Integrating eye care services into universal health coverage will ensure affordable healthcare services for PLWD and prevent avoidable blindness due to DR.

PLWD reported that they sometimes prioritised their businesses and work over eye health because they did not have enough time. Integrating DR screening services with DM care will allow PLWD to access DR services while receiving DM care ^7,17^ hence reducing the time needed to seek other healthcare services.

Some participants claimed to have never utilised diabetic eye care services before because they were not referred by their primary healthcare service provider, while others went for eye care services after being referred from primary healthcare facilities. Referral from primary healthcare facilities plays a great role in PLWDs’ access to timely eye care.^12^

The participants in the present study reported that they sought eye care services after experiencing symptoms such as blurred vision. This has also been reported in previous studies in which PLWD perceived that they only needed eye care after they began to experience a problem with their vision^12^. PLWD should be educated about the importance of DR screening so that they can report earlier, before they start to experience vision deterioration and reach the hospital at a point where there is a good prognosis after receiving treatment.

We recommend strengthening health promotion activities and educating PLWD that DM causes vision loss and the importance of attending DR screening as per their Doctor’s recommendations, but also educating PLWD about the safety of treatment services of DR will help PLWD adhere to diabetic eye care services, and also involving Physicians and other primary healthcare providers in improving referral pathways for PLWD to eye healthcare services.

In this study we ensured the trustworthiness of research findings by member checking where we asked some of the participants if the results portrayed what they said during interviews, also data were mainly analysed an Optometrist (HN) with training in qualitative research, and an Associate Professor of public health (DK), peer debriefing was also done by consulting peers who are knowledgeable on qualitative research methods.

This study had several limitations which are; only one Hospital in an urban area was included and the views of people attending that hospital might be very different from those of other people in rural areas, we also sampled a population of PLWDs who are already attending eye care services, these limitations were mitigated by including participants from different regions of the country attending the tertiary Hospital.

## Conclusion

In this study, we explored the factors influencing access to diabetic eye care among PLWD, the costs of eye care services, not knowing that DM can affect eyes, fear and competing priorities emerged as barriers to accessing care while referral from primary healthcare workers, eye symptoms, knowing that diabetes can affect eyes, living close to the facility offering eye care services, influence from close relatives, and having health insurance were reported as facilitators to attending diabetic eye care services.

Educating PLWD about the importance of diabetic eye care while working toward overcoming the barriers to accessing DR eye care will help PLWD access timely diabetic eye care and prevent blindness due to DR.

## Data Availability

All data produced in the present study are available upon reasonable request to the authors

## References

1. Associates Y and. Global Prevalence and Major Risk Factors of Diabetic Retinopathy. Diabetes Care. 2012;35:556–64.

2. Burgess PI, MacCormick IJC, Harding SP, Bastawrous A, Beare NAV, Garner P. Epidemiology of diabetic retinopathy and maculopathy in Africa: A systematic review. Diabet Med. 2013;30(4):399–412.

3. Zheng Y, He M, Congdon N. The worldwide epidemic of diabetic retinopathy. Indian J Ophthalmol. 2012;60(5):428–31.

4. Cleland CR, Burton MJ, Hall C, Hall A, Courtright P, Makupa WU, et al. Diabetic retinopathy in Tanzania: Prevalence and risk factors at entry into a regional screening programme. Trop Med Int Heal. 2016;21(3):417–26.

5. Mtuya C, Cleland CR, Philippin H, Paulo K, Njau B, Makupa WU, et al. Reasons for poor follow-up of diabetic retinopathy patients after screening in Tanzania: A cross-sectional study. BMC Ophthalmol [Internet]. 2016;16(1):1–7. Available from: 10.1186/s12886-016-0288-z

6. Mumba M, Hall A, Lewallen S. Compliance with eye screening examinations among diabetic patients at a Tanzanian referral hospital. Ophthalmic Epidemiol. 2007;14(5):306–10.

7. Omar FJ, Sheeladevi S, Rani PK, Ning G, Kabona G. Evaluating the effectiveness of opportunistic eye screening model for people with Diabetes attending Diabetes clinic at Mnazi Mmoja hospital, Zanzibar. BMC Ophthalmol. 2014;14(1):1–7.

8. Michie S, Stralen MM Van, West R. The behaviour change wheelLJ: A new method for characterising and designing behaviour change interventions The behaviour change wheelLJ: A new method for characterising and designing behaviour change interventions. Implement Sci. 2011;6(42):1–12.

9. Timlin D, Mccormack JM, Simpson EEA. Using the COM-B model to identify barriers and facilitators towards adoption of a diet associated with cognitive function (MIND diet). Public Health Nutr. 2020;24(7):1657–70.

10. Madhani A, Finlay KA. Using the COM--B model to characterize the barriers and facilitators of pre--exposure prophylaxis (PrEP) uptake in men who have sex with men. Br J Health Psychol. 2022;27:1330–53.

11. Ford BK, Angell B, White AJR, Duong A, Hiidome S, Keay L. Experiences of Patients With Diabetes Attending a Publicly Funded Eye Care Pathway in Western Sydney: A Qualitative Study. J Patient Exp. 2021;8:1–12.

12. Mwangi N, Macleod D, Gichuhi S, Muthami L, Moorman C, Bascaran C, et al. Predictors of uptake of eye examination in people living with diabetes mellitus in three counties of Kenya. Trop Med Heal. 2017;45(41):1–10.

13. Lake AJ, Browne JL, Rees G, Speight J. What factors influence uptake of retinal screening among young adults with type 2 diabetes? A qualitative study informed by the theoretical domains framework. J Diabetes Complications [Internet]. 2017;31(6):997–1006. Available from: 10.1016/j.jdiacomp.2017.02.020

14. Fairless E, Nwanyanwu K. Barriers to and Facilitators of Diabetic Retinopathy Screening Utilization in a High-risk Population. J Racial Ethn Heal Disparities. 2020;6(6):1–12.

15. ‘Ofanoa M, Aitip B, Ram K, Dalmia P, Pal M, Nosa V, et al. A qualitative study of patient perspectives of diabetes and diabetic retinopathy services in Vanuatu. Heal Promot J Aust. 2022;33(1):289–96.

16. Hall CE, Hall AB, Kok G, Mallya J, Courtright P. A needs assessment of people living with diabetes and diabetic retinopathy. BMC Res Notes. 2016;9(1):1–14.

17. Mwangi N, Bascaran C, Gichuhi S, Kipturgo M, Manyara L, Macleod D, et al. Rationale for integration of services for diabetes mellitus and diabetic retinopathy in Kenya. Springer Nat. 2022;36:4–11.

